# Blood Levels of NLRP3 Inflammasome Components is Associated with Obsessive Compulsive Disorder

**DOI:** 10.1101/2020.01.30.20019646

**Authors:** Melike Tetik, Nese Direk, Betul Onder, Cansu Aykac, Burcu Ekinci, Tutku Yaras, Aykut Kuruoglu, Cagatay Ermis, Tunc Alkin, Yavuz Oktay

**Author notes:** **CORRESPONDING AUTHOR:** Asst.Prof.Dr. Yavuz Oktay, Izmir Biomedicine and Genome Center, Dokuz Eylul University, Faculty of Medicine, Department of Medical Biology, 35340 Balcova, Izmir, Turkey.

## Abstract

**Background:** Inflammation has a well-known role in the pathogenesis of a range of neuropsychiatric disorders such as major depressive disorder and schizophrenia. Previous studies provided evidence regarding its possible involvement in the etiology of obsessive-compulsive disorder (OCD). However, mechanisms explaining the association of inflammation with OCD are lacking. The NLRP3 inflammasome complex initiates and mediates inflammatory response, which explain one of the most important key mechanisms behind inflammatory response. In this study, we aimed to determine a possible association between NLRP3 inflammasome and OCD, and to evaluate its relationship with clinical features.

**Methods:** This case-control study included 103 participants (51 OCD and 52 healthy controls). OCD patients were diagnosed using DSM-IV criteria. All participants were evaluated by psychiatric inventories, i.e. Yale Brown Obsessive Compulsive Scale, Hamilton Depression Scale, and Hewitt Multidimensional Perfectionism Scale. Peripheral blood mononuclear cells (PBMCs) were isolated from fresh blood samples. RNA and protein were extracted from PBMCs and expression of NLRP3 inflammasome components were evaluated by quantitative real-time PCR (qPCR) and Western blotting. Serum IL-1beta and IL-18 cytokine levels were determined by ELISA.

**Results:** NEK7 and CASP1 mRNA levels were significantly higher in OCD patients, compared to controls. Pro-Caspase-1 protein levels were elevated, as well. Regression analysis showed that NEK7 mRNA and Caspase-1 protein levels can differentiate OCD and healthy control groups.

**Discussion:** pro-Caspase-1 mRNA and protein levels in PBMCs, as well as NEK7 mRNA levels are potential biomarker candidates in OCD. Our results may prompt research into possible role of NLRP3 inflammasome in OCD etiology.

## INTRODUCTION

Obsessive Compulsive Disorder (OCD) is characterized by repetitive and involuntary thoughts and behaviors that disrupt daily activities of the affected person. OCD is a symptomatically heterogeneous disorder with several factors involved in etiology. One of these factors is inflammation, which is a non-specific etiological factor for OCD (1). Inflammation in OCD has been studied constantly. Generally, peripheral levels of pro-inflammatory and anti-inflammatory cytokines were used as markers of inflammation in OCD studies, which provide limited information about the role of inflammation as a pathophysiological factor of OCD.

Inflammasomes are the sensors of various stress stimuli and mediators of inflammatory response. They are oligomeric structures composed of Nucleotide-binding oligomerization domain (NOD)-like receptor (NLR) proteins. Nucleotide-binding oligomerization domain, Leucine rich Repeat (LRR) and Pyrin domain (PYD) containing protein 3 (NLRP3) inflammasome is formed by the interaction of NLRP3, pro-Caspase-1 and Apoptosis-associated speck-like protein containing a CARD (PYCARD/ASC) proteins. It has been related to neuropsychiatric conditions such as Major Depressive Disorder (MDD), Bipolar Disorder, Schizophrenia, Parkinson’s Disease and Multiple Sclerosis (1–6). Upon stress stimuli, NLRP3 protein is activated to form an oligomeric complex in the cells that is known as inflammasome complex. Active inflammasome complex cleaves pro-Caspase-1 to yield active Caspase-1 that processes cytokines such as IL-1β, IL-18 and IL-33.

The Nima-Related Kinase 7 (NEK7) was recently identified as a regulator of NLRP3 inflammasome activity. It was previously known to organize microtubule formation and mitosis. Inflammasome-activating signals cause potassium efflux and therefore the stimulation of NEK7, which leads NLRP3 oligomerization and speckle formation (7–9). NEK7 is required for activation of NLRP3 inflammasome and it is specific to NLRP3 as it has no function on the activation of NLRC4 and AIM2 (7,9).

There are only few studies in which an association of inflammasomes and psychiatric disorders including psychosis, bipolar disorder and MDD has been tested. However, such relationship has not been tested for OCD. Considering the increasing evidence that suggests involvement of neuroinflammation in OCD pathogenesis, we hypothesized that NLRP3 inflammasome complex might be related with OCD. Therefore, we compared mRNA and protein levels of NLRP3 inflammasome components and NEK7 between OCD patients and healthy controls. We also investigated the possible association between NLRP3 inflammasome components and clinical features of OCD, such as severity, co-existing anxiety, depression, and perfectionism.

## Methods and Materials

### Study Design

#### Study Population

The study was performed at the Department of Psychiatry, Dokuz Eylul University Hospital between February and November 2018. All protocols and methods were approved by the Dokuz Eylul University Clinical Studies Ethical Committee (2017/08-06, Protocol number 378-SBKAEK). In total, 51 patients with OCD (aged 18 to 45 years) and 52 healthy controls were enrolled in the study. Oral and written informed consents were obtained from all participants. Healthy controls were age and sex-matched participants with no history of a chronic disease or psychiatric disorder. Participants with an inflammatory disorder such as rheumatoid arthritis, autoimmune disorders (i.e. Hashimoto’s thyroiditis) and those on corticosteroid, statin, antihypertensive or non-steroid anti-inflammatory treatments were excluded from the current study. If a participant used an anti-inflammatory medication in the past five days, blood collection and psychiatric evaluations were postponed. Additionally, people with any sign of infection within one week prior to blood collection were not included in this study.

#### Clinical evaluation

Sociodemographic data was collected. The Structured Clinical Interview for DSM-IV Axis I Disorders (SCID-I) was applied to confirm OCD diagnosis and to assess the presence of other psychiatric disorders. Yale-Brown Obsession and Compulsion Scale (Y-BOCS) was used to identify the severity of obsessive and compulsive symptoms (10). Severity of depressive symptoms and level of perfectionism were evaluated using Hamilton Depression Rating Scale (HAM-D) and Hewitt’s Multidimensional Perfectionism Scale (MPS), respectively (11,12).

#### Molecular Analyses of Inflammatory Markers

##### Peripheral Blood Mononuclear Cell (PBMC) Isolation

For molecular studies, fasting peripheral venous blood samples (16.5 ml) were collected between 9:00 and 11:00 a.m.. PBMCs were isolated from fresh blood samples (6 ml) by adding 1:1 vol:vol Phosphate Buffered Saline (PBS) and overlaying on 3 ml lymphocyte separation medium (Lonza, Cat#BE17-829E), followed by centrifugation (Eppendorf 5810 R) at 2200 rpm for 25 minutes. Isolated PBMCs were washed in PBS at 1500 rpm for 10 minutes. In total, 500 μl TRIzol (ThermoFisher) was added to pellets to preserve RNA integrity. 4 aliquots/sample were stored at −80 °C until RNA and protein isolation.

##### RNA Isolation and cDNA Synthesis

Aliquots in TRIzol were thawed on ice and 1:1 vol:vol EtOH (100%) was added. RNA extraction was performed using Direct-zol RNA Miniprep Plus kit (Zymo, Cat# R2072) according to the manufacturer’s protocol and stored at −80 °C until cDNA synthesis. RNA purity and quantity were assessed using Nanodrop 2100c spectrophotometer (Thermo Scientific). OD_260/280_ and OD_260/230_ ratios were calculated and samples with 1.9-2.0 and >1.5, respectively and were used for cDNA synthesis. One μg RNA was converted to cDNA using First Strand cDNA Synthesis kit (ProtoScript Cat# E6300).

#### Quantitative Real-Time PCR (qPCR)

mRNA expression levels of NLRP3, PYCARD, CASP1 and NEK7 were determined using GoTaq qPCR Master Mix (Promega, Cat# A6001) on LightCycler480 (Roche). GAPDH was used as housekeeping gene for normalization and relative expression of each gene was calculated according to 2^-Ct^ method. Primer pairs used for each gene are: NEK7_forw 5’- TTTACACTCCTGACAGCG -3’, NEK7_rev 5’-GCAACAGGAACTTTAGAACT -3’, Caspase-1_forw 5’- CTCAGGCTCAGAAGGGAATG-3’, Caspase-1_rev 5’-CGCTGTACCCCAGATTTTGT -3’, NLRP3_forw 5’- GCAGCAAACTGGAAAGGAAG-3’, NLRP3_rev 5’- CTTCTCTGATGAGGCCCAAG- 3’, ASC_forw 5’-AGTTTCACACCAGCCTGGAA -3’, ASC_rev 5’- TTTTCAAGCTGGCTTTTCGT-3’, GAPDH_forw 5’- ACCACAGTCCATGCCATCAC -3’, GAPDH_rev 5’- TCCACCCTGTTGCTGTA -3’. Each reaction was performed in triplicates and melting-curve analysis was performed for each primer pair to ensure specificity.

#### Protein Extraction from PBMCs and Western Blotting

Chloroform was added on TRIzol-protected samples at 1:5 vol:vol ratio. Following centrifugation and EtOH wash, proteins were precipitated with 0.3 M guanidium hydrochloride. After washing, pellets were dissolved in 1% SDS solution. Protein concentration was measured using BCA Assay Kit (Thermo, Cat#: 23227). Equal amount of protein (30 μg) was mixed with SDS-sample buffer and boiled for 5 min to denature proteins. Proteins were separated on SDS-Polyacrylamide gel in Tris-Glycine Buffer for 2 hours and transferred to PVDF membrane at 250 mA for 2 hours. Membrane with transferred proteins was blocked with 5% fat-free milk in 1X TBS (Tris-buffered saline) solution. Optimal incubation times for each primary antibody was determined and followed by incubation with the appropriate horse-radish-peroxidase (HRP)-conjugated secondary antibody. Antibodies used for each protein are: anti-CASP1 (abcam Cat# ab1872), anti-PYCARD (AG, Cat# 25B-0006-C), anti-NEK7 (abcam, Cat#ab13351), anti-NLRP3 (AG, Cat# 20B-OO14-C), anti-beta-actin (abcam, Cat# ab6227). Finally, membranes were incubated with Luminata Forte Western HRP Substrate (Luminata, WBLUF0500) and chemiluminescence was measured on imaging system (Vilber Lourmat, ECX-F20L). Image analysis was performed using the on-site software and protein levels were calculated with reference to beta-actin.

#### Enzyme-linked Immunosorbent Assay (ELISA) Analysis

ELISA analysis was performed on serum samples isolated from blood collected into yellow-cap gel-bottom tubes. Following a 30-minute incubation at room temperature, samples were centrifuged at 4000 rpm (2850 g) for 5 minutes and 500 μl aliquots were stored at −80 °C until measurement. ELISA was performed according to the manufacturer’s instructions for each protein: IL-1beta (Thermo Scientific, Invitrogen, Cat# BMS224-2) and IL-18 (Thermo Scientific, Invitrogen, Cat# BMS267NST). 50 μl serum sample from each participant was used and absorbance at 450 nm – 620 nm was measured (Varioskan, Thermo). Standard curves were used to determine IL-1beta and IL-18 concentration of each sample.

### Statistical analysis

IBM SPSS Statistics v. 22.0 was used for statistical analyses. We first tested the distribution of the continuous variables using histograms with normality curves. Variables with non-normal distribution were transformed using natural logarithm (LN). We excluded outliers (n=3) when analyzing inflammatory parameters that were assessed with Western blotting. Descriptive analyses were done using a chi-square or t-test.

Associations between inflammatory parameters measured with qPCR, Western blotting and ELISA and OCD status were evaluated with logistic regression analyses. In the first model, we adjusted the analyses for age, gender and duration of education. In the second model, Hewitt’s MPS total score were entered into the model. In these analyses, LN-transformed values of inflammatory parameters were used, and results were given as back-transformed values.

In the OCD group, we tested possible predictors of the inflammatory markers using a linear regression analysis. In these analyses, age, gender, educational attainment, Y-BOCS score, total duration since OCD diagnosis, presence of OCD medication, Hewitt’s MPS total score and HAM-D total scores were used as predictors. Again, back-transformed results were presented.

## Results

Characteristics of study population are presented in Table 1. A total of 103 volunteers participated in the study, 51 of them were diagnosed with OCD and 52 volunteers had no history of psychiatric disorders. The mean age of OCD patients and healthy controls were 30.41 ± 8.90 and 30.56 ± 7.89, respectively. In total 60.8% (n=31) of the OCD group and 59.6% (n=31) of the healthy controls were female.

**Table 1.**
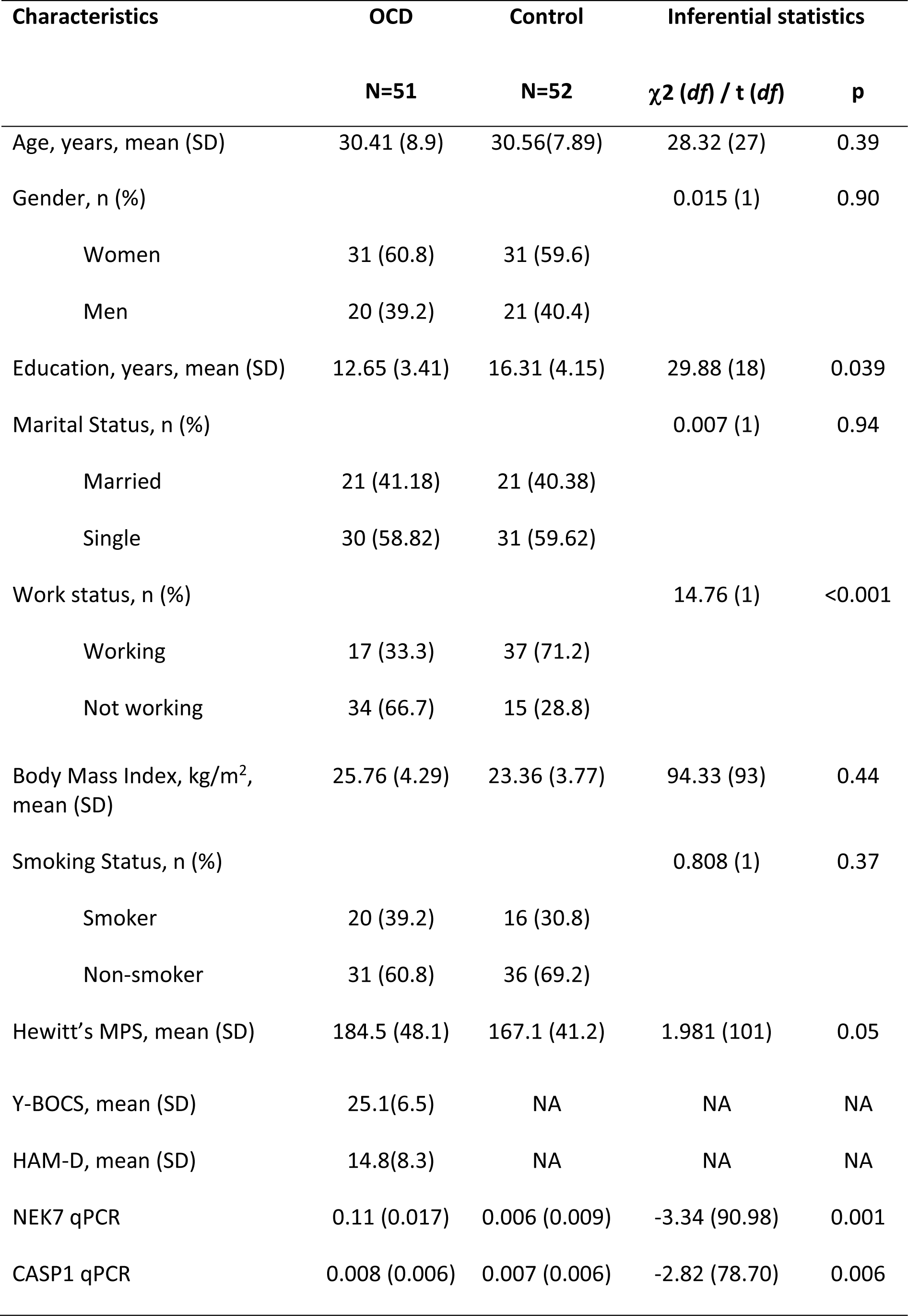
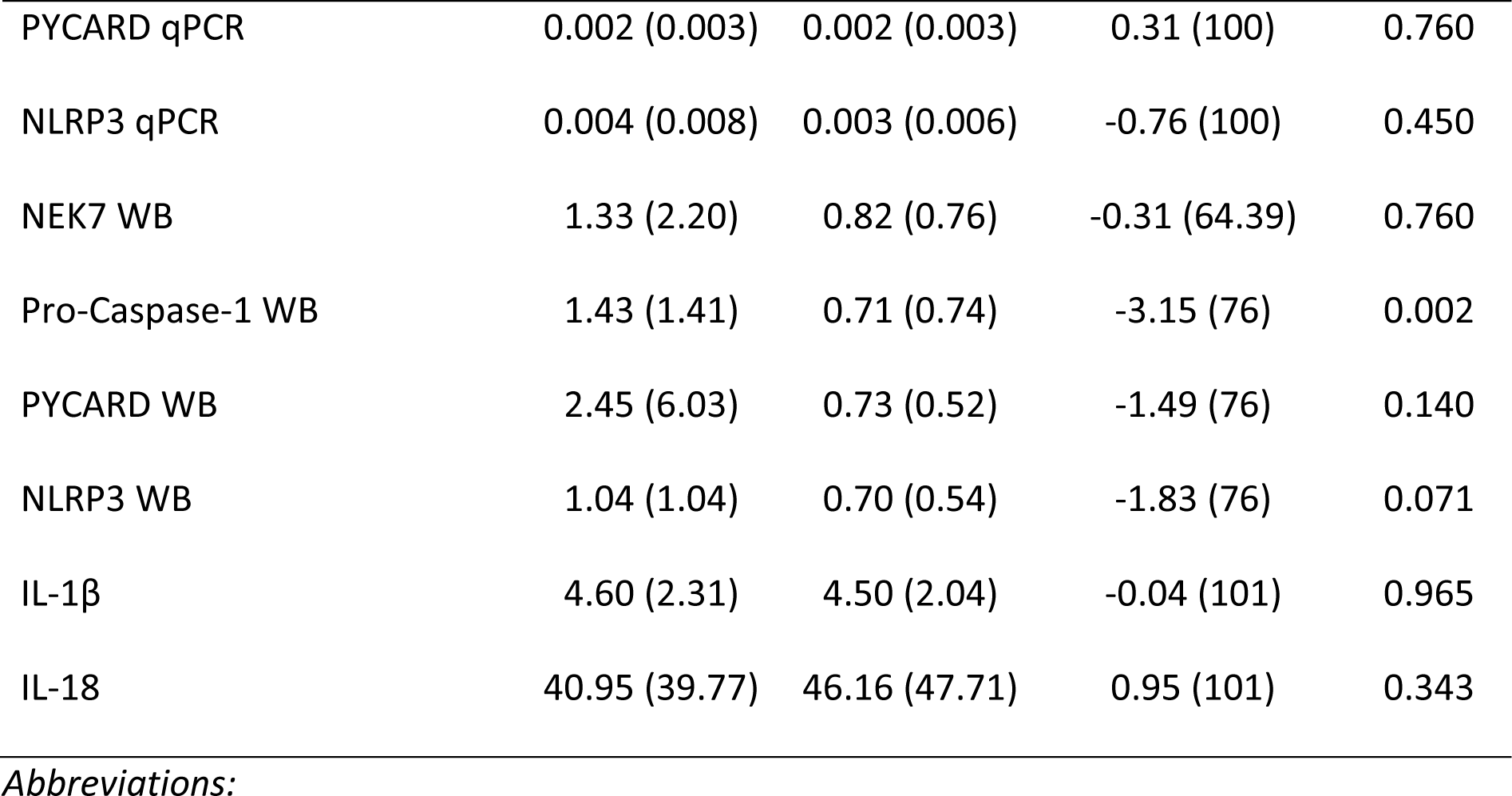
Characteristics of the study population.

In OCD group, 21.6% (n=11) had comorbid dysthymia, 17.6% (n=9) had MDD, 17.6% (n=9) had generalized anxiety disorder, 9.8% (n=5) had specific phobia, 5.9% (n=3) had panic disorder, 2% (n=1) had post-traumatic stress disorder. Also, 9.8% (n=5) of OCD group had a history of MDD and 3.9% (n=2) had a history of post-traumatic stress disorder. In total, 45.1% (n=23) of OCD patients were on antidepressant treatment.

Hewitt’s MPS scores were higher in OCD patients compared to healthy controls whereas the difference was not statistically significant (184.5 ± 48.1 vs. 167.1 ± 41.2, p=0.05). Mean YBOC-S score was 25.1 ± 6.5 and mean HAM-D score was 14.8 ± 8.3 in OCD group (Table 1). NEK7 mRNA (p=0.001), CASP1 mRNA (p=0.003) and pro-Caspase-1 protein levels (p=0.002) were higher in OCD patients compared to healthy controls (Table 1).

A series of logistic regression analyses were performed to test the association of mRNA and protein levels of NLRP3, Caspase-1, PYCARD and NEK7 with OCD status. Final models in which analyses were adjusted for age, sex, length of educational attainment, Hewitt’s MPS scores indicated that NEK7 mRNA and pro-Caspase-1 protein levels are associated with OCD (Table 2). On the other hand, there was no association between NLRP3 or PYCARD mRNA/protein levels with OCD.

**Table 2.** Regression Analyses.

|  | **OR** | **95 % CI** | ***P*** |
| --- | --- | --- | --- |
| NEK7 qPCR |  |  |  |
| Model 1 | 1.89 | 0.348-0.970 | .003 |
| Model 2 | 1.63 | 0.387-0.970 | .036 |
| CASP1 qPCR |  |  |  |
| Model 1 | 0.61 | 0.418-0.884 | .009 |
| Model 2 | 0.70 | 0.463-1.061 | .093 |
| PYCARD qPCR |  |  |  |
| Model 1 | 1.06 | 0.746-1.498 | .755 |
| Model 2 | 1.16 | 1.162-1.739 | .467 |
| NLRP3 qPCR |  |  |  |
| Model 1 | 1.13 | 0.822-1.560 | .446 |
| Model 2 | 1.16 | 0.805-1.674 | .424 |
| NEK7 WB |  |  |  |
| Model 1 | 1.07 | 0.707-1.614 | .754 |
| Model 2 | 0.98 | 0.625-1.540 | .934 |
| Pro-Caspase-1 WB |  |  |  |
| Model 1 | 2.10 | 1.252-3.533 | .005 |
| Model 2 | 2.12 | 1.190-3.791 | .011 |
| PYCARD WB |  |  |  |
| Model 1 | 1.35 | 0.900-2.034 | .147 |
| Model 2 | 1.25 | 0.811-1.923 | .312 |
| NLRP3 WB |  |  |  |
| Model 1 | 1.50 | .955-2.341 | .079 |
| Model 2 | 1.50 | .925-2.428 | .101 |

Linear regression analysis was conducted to test for possible predictors of NLRP3 inflammasome components in OCD group. There was no significant predictor of mRNA levels in OCD group. OCD patients who are on medication had lower NLRP3 protein levels (p=0.03). Similarly, PYCARD protein levels were lower in OCD patients on treatment. In the same model, age and Y-BOCS score were negatively associated with PYCARD protein levels in OCD patients. There was no significant predictor of NEK7, pro-Caspase-1, IL-1beta or IL-18 levels in OCD group (Supplementary Table 1).

We also tested possible association between NLRP3 inflammasome and depression levels among OCD patients. OCD patients were grouped according to the commonly used cut-off score for HAM-D scores (i.e. <18 and ≥18). mRNA levels of NLRP3, PYCARD, Caspase-1 and NEK7 were not related to depression status in OCD group. On the other hand, NEK7 protein levels were significantly different between the two groups (p=0.03). There was no association of IL-1beta or IL-18 levels with depression status in OCD group (Supplementary Table 2).

## Discussion

To our knowledge, this is the first study exploring and indicating an evidence for an association between NLRP3 inflammasome pathway and OCD. Caspase-1 mRNA and pro-Caspase-1 protein levels, and NEK7 mRNA levels were higher in OCD patients compared to healthy controls. Also, NEK7 mRNA and pro-Caspase-1 protein levels were related to OCD status independent of confounders.

We did not observe any significant differences in NLRP3 and PYCARD mRNA/protein expression levels between OCD and healthy controls. It should be kept in mind that activation of biological processes usually occurs by upregulation or activation of a limited number of molecules that tend to be rate-limiting, rather than upregulation of most or all components. It is possible that increased NEK7 mRNA levels in OCD patients reflect altered epigenetic states that lead to increased NEK7 transcription and/or mRNA stability. As an upstream regulator, NEK7 was shown to be critical for transmission of activation signals to NLRP3 inflammasome and it could be speculated that its levels determine the sensitivity or strength of inflammasome activation. In other words, higher NEK7 levels may make cells more sensitive to even low levels of inflammasome-activating signals. Coupled with increased baseline stress levels, as in neuropsychiatric disorders, increased sensitivity to inflammatory agents may cause pathologic activation of NLRP3 and lead to diseases (13). Further studies where cells isolated from OCD patients and exposed to various stimuli (e.g. LPS) would help to test this hypothesis. Epidemiological studies where NLRP3-activating stimuli are investigated may also help to establish the causal relationship between NLRP3 inflammasome and OCD.

On the other hand, NEK7 has recently been shown to mark GABAergic parvalbumin-positive (Pv+) cortical interneurons and control their wiring (14). This specific subset of interneurons provide feedback to pyramidal neurons and are required for the oscillatory activity in the gamma-frequency (15) and their maturation parallels the refinement of cortical circuits (16). Defects in Pv+ cortical interneurons has been associated with sociability defects, similar to those in autism spectrum disorder and schizophrenia (17). Barnes et al. showed that disruption of the metabotropic glutamate receptor 5 (mGluR5) in Pv+ interneurons caused compulsive-like repetitive behaviour and increased prepulse inhibition (16). Therefore, it would be intriguing to test whether increased NEK7 expression is also present in Pv+ interneurons of OCD post-mortem brains. We cannot rule out this alternative explanation, however, increased CASP1 expression in addition to NEK7 suggests involvement of the NLRP3 inflammasome.

A limitation of our study is the high rate of psychiatric medicine use among our OCD patients (n=40/51, %88), which may skew the results. Accordingly, fluoxetine use was shown to suppress activation of NLRP3 activation (18). In another study, nine different antidepressants was shown to reduce IL-1β and IL-18 levels in depression patients, both *in vivo* and *in vitro* (19). Moreover, the authors suggested use of IL-1β and IL-18, as well as NLRP3 inflammasome levels, as biomarkers of response to anti-depressants in major depression patients. However, no such association is currently known between anti-depressant use and the levels of NEK7 or Caspase-1. It is possible that our observation of upregulation of only some components of NLRP3 inflammasome in OCD patients may be at least partially explained by suppression of the other components by long-term anti-depressant use.

Another confounding factor may be inclusion of patients with depression diagnosis, as NLRP3 activation has been linked to major depressive disorder (MDD) (4). However, currently there is no knowledge on such an association between NEK7 and MDD. Therefore, increased NEK7 expression in OCD patients cannot simply be explained by MDD comorbidity. Similarly, we assessed possible interference of depression on increased pro-Caspase-1 levels by comparing patients with HAM-D score <18 to those with HAM-D score ≥18, which is close to syndromal level. Lack of any difference between the two groups in terms of mRNA or protein levels suggest that increased pro-Caspase-1 levels in OCD patients is independent of depression comorbidity.

As is true of all complex human phenotypes, measurement of inflammation-associated molecules in humans is affected by a multitude of factors. Despite this difficulty, our observation of two different components of NLRP3 pathway being significantly associated with OCD in a relatively small cohort is remarkable: NEK7 at mRNA level and pro-Caspase-1 at protein level. While the other components of the pathway failed to show any such association, there was a common trend towards upregulation that did not reach statistical significance. On the other hand, the discrepancies between qPCR and Western blot measurements are widely reported in literature across disease states and tissues, reflecting a fundamental aspect of gene regulation. While Western blotting is a semi-quantitative method, despite measuring the effector molecules in cells/body, qPCR is a more quantitative method preferred in a wide range of diseases where accurate quantitation is critical, such as viral load determination in hepatitis and HIV., cancer diagnosis and follow-up etc. (20). Therefore, discrepancies between results obtained by qPCR and Western blotting should be interpreted accordingly.

Role of inflammation is much less clear in the pathogenesis of OCD compared to other neuropsychiatric diseases (e.g. MDD) and has largely been limited to measurement of cytokines in blood or isolated macrophages. By providing evidence that suggests possible involvement of NLRP3 pathway alterations in OCD pathogenesis, our findings may lead to new avenues of research into OCD-inflammation association. At the very least, addition of inflammasome components to the list of frequently measured inflammation-associated molecules can be expected. Particularly NEK7 and CASP1 mRNA measurement from PBMCs may provide highly quantitative biomarkers that can help diagnosis, monitor disease progression, measure response to treatment etc. Studies on bigger cohorts will test our findings and will ideally include drug-naive and depression-free patients. Mechanistic studies will also help establish the causal relationship between NLRP3 inflammasome and OCD.

## Data Availability

Data availability is upon request.

## Acknowledgements

This study was supported by TUBİTAK project 217S128 (T. Alkın and Y. Oktay).

